# Societal feedback induces complex and chaotic dynamics in endemic infectious diseases

**DOI:** 10.1101/2023.05.25.23290509

**Authors:** Joel Wagner, Simon Bauer, Sebastian Contreras, Luk Fleddermann, Ulrich Parlitz, Viola Priesemann

## Abstract

Classically, endemic diseases are expected to display relatively stable, predictable infection dynamics. Indeed, diseases like influenza show yearly recurring infection waves that can be anticipated accurately enough to develop and distribute new vaccines. In contrast, newly-emerging diseases may cause more complex, unpredictable dynamics, like COVID-19 has demonstrated. Here we show that complex infection dynamics can also occur in the endemic state of seasonal diseases when including human behaviour. We implement human behaviour as a feedback between incidence and disease mitigation and study the system as an *epidemiological oscillator* driven by seasonality. When behaviour and seasonality have a comparable impact, we find a rich structure in parameter and state space with Arnold tongues, co-existing attractors, and chaos. Moreover, we demonstrate that if a disease requires active mitigation, balancing costs of mitigation and infections can lead societies right into this complex regime. We observe indications of this when comparing past COVID-19 and influenza data to model simulations. Our results challenge the intuition that endemicity implies predictability and seasonal waves, and show that complex dynamics can dominate even in the endemic phase.

---

Infectious diseases have always accompanied humans and strongly impacted societies. Some of them emerge and are then eliminated again. Others come to stay, and their repeated outbreaks or steady prevalence may require structured mitigation [1]. Understanding the fundamental mechanics governing their spread and mitigation enables us to predict their short and long-term dynamics (“waves” of incidence) [2, 3], which is the basis to develop adequate policies [4, 5].

But predictability varies. On one end of the spectrum are e.g. seasonal viruses, like seasonal influenza or respiratory syncytial viruses (RSV), whose waves follow clear yearly patterns [6, 7]. These have reached an *endemic* steady-state: Partial immunity in the population has established a steady prevalence, potentially modulated by seasonality. On the other end of predictability are diseases such as Ebola or Marburg virus disease that stochastically emerge and re-emerge (e.g., from environmental reservoirs). These pose an immense threat to public health, calling for rapid elimination. Their inherent threat to health prevents them from becoming endemic and thus large fractions of the population remain susceptible. On this scale, COVID-19 has provided an important middle ground: It was lethal enough to have required significant mitigation, but was not eliminated. In its first years, the combination of intermittent mitigation, new variants, and seasonality led to complex, potentially chaotic dynamics with off-seasonal waves and strong differences between countries [4], thereby challenging the prediction of subsequent waves [8–10]. Was this complex dynamics solely an expression of the pandemic (initial) phase of COVID-19, or is it a general property of infectious diseases that can also emerge in the endemic state?

Complexity and unpredictability in wave patterns arise either from the stochasticity of contagion, the emergence of new variants, or variable mitigation measures (human behaviour). The impact of these factors are classically studied in agent-based or compartmental SIR(S)-like models [11–13]. In compartmental SIRS models, the population is split into disjoint compartments according to their disease status. Susceptible individuals (S) get infected by infectious individuals (I), eventually recover (R) and stay immune until they become susceptible again (S). Due to the waning of immunity, diseases persist at an equilibrium level, known as the endemic equilibrium. Such SIRS models have been adapted and extended to study the effects of either seasonality or human behavioural adaptation (mitigation) on their own [9, 10, 14–17]. However, these two components may interact, and their combined effect might give rise to complex dynamics - the extend of which is unclear.

It is a core challenge to incorporate human behaviour into disease models [10, 11, 14, 16]. One natural approach is to assume that high incidences are perceived as increased *hazard h*. People subsequently mitigate the spread by reducing their contacts, quarantining, wearing mask or adopting other health-protective behaviour—voluntarily or due to governmental mandates [10, 17]. Such a feedback mechanism can turn the endemic equilibrium unstable through a supercritical Hopf bifurcation, leading to behaviour-induced oscillations [16]: Contact reduction due to increased hazard mitigates the spread, lowering the incidence and perceived hazard, which ultimately triggers a rebound wave. Thereby, periodic waves can emerge.

Periodicity also arises from the seasonal variation of the spreading rate. If such seasonality is applied to disease spread with intrinsic behaviour-induced oscillations, one can view this system as an externally driven *epidemiological oscillator*. From dynamical systems theory, such driven oscillators are known to display very complex steady-state dynamics [18–20], including narrow regimes of phase-locking, chaos and coexisting attractors. Hence, one might find similar complexity for seasonal endemic diseases with far-reaching consequences for forecasts, predictability and policy planning: Chaotic dynamics, together with already small changes in human behaviour, infectiousness or initial conditions, could vastly alter the wave patterns, limiting the predictive power of forecasts. However, it remains unclear whether and under which conditions such complex dynamics can arise in epidemiology.

Here, we show that the entanglement of seasonality and mitigation gives indeed rise to a vast variety of endemic disease states. These range from the classical seasonal waves to strongly patterned regimes in parameter space where phase-locking, multi-stability, and chaotic dynamics occur. Moreover, we demonstrate that balancing costs of mitigation against costs of infections can lead societies right into the complex regimes. Comparing past data of seasonal influenza and COVID-19 waves to simulations provides evidence that this balancing can explain their vastly differing dynamics. This demonstrates that complex dynamics and limited predictability that characterised e.g. the initial phases of COVID-19 are not limited to the initial phases of a disease, but can, contrary to classical belief, extend into endemicity.

## Model overview

To investigate the interplay between seasonality and human behavioural feedback on disease spread, we used a modified susceptible-infectious-recovered-susceptible (SIRS) compartmental model, incorporating seasonality and behavioural feedback for mitigation (m) (SIRSsm model, Fig. 1a). Seasonality is represented by a periodic driving *s*(*t*) = 1 + *a* cos(*ωt*), which modulates the basic reproduction number *R*_0_ with a yearly cycle. The behavioural feedback accounts for mitigation *m*(*h*) (voluntary and mandatory) that reduces the spreading rate of the disease, depending on perceived hazard *h* (see below). Altogether, the effective reproduction number *R*^eff^(*t*) reads

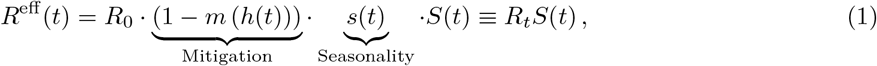

where 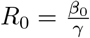 is expressed as ratio between the spreading rate *β*_0_ and recovery rate *γ* (see Tab. 1 for all variables and default values); *S*(*t*) is the susceptible fraction of the population. *R*_*t*_ represents the effective reproduction number without accounting for immunity and is thus a measure for the contact levels and the infectiousness of encounters.

**Figure 1:**
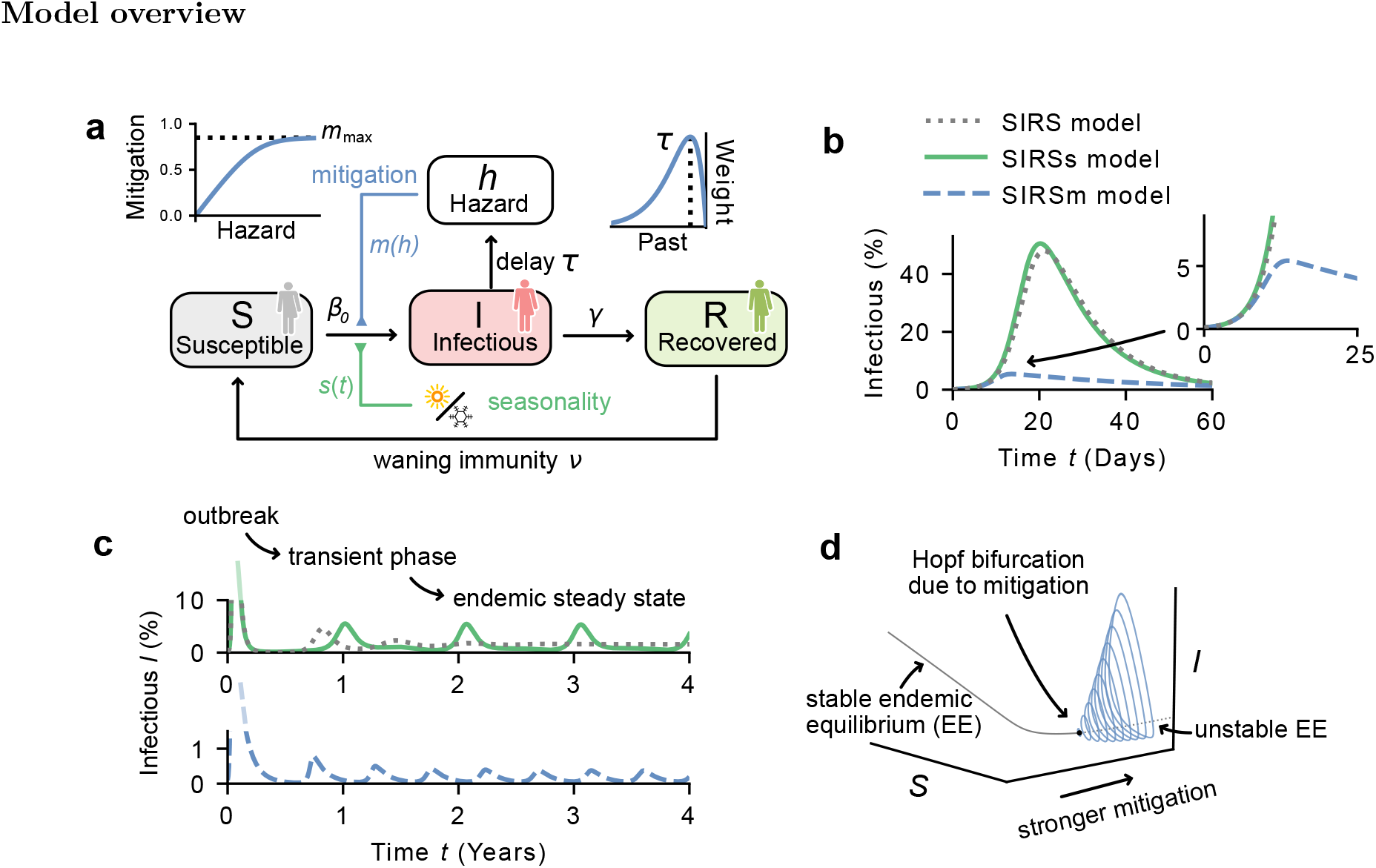
Model overview. **a)** The SIRSsm model: An extended SIRS model including periodic seasonality (*s*, green) and mitigation from behavioural feedback (*m*, blue). The population is divided into compartments according to their disease status: Susceptible (S), infectious (I) or recovered (R). Mitigation *m* increases with hazard *h* and saturates at a maximum mitigation level of *m*_max_. Hazard *h* is calculated as the convolution of the infectious compartment *I* with a delaying kernel of mitigation delay *τ* (peak of the kernel). Yearly seasonality *s*(*t*) periodically modulates the infection rate. **b)** For a novel disease outbreak, classic SIRS models (with or without seasonality *s*, resp. green and grey) feature initial exponential growth in incidence, slowed down and eventually stopped only by immunity. In contrast, with behavioural feedback, the incidence is kept considerably lower due to mitigation (blue, see inset). **c)** Following the initial outbreak and a transient phase, incidence levels settle into their endemic steady state. In SIRS models without feedback, this equilibrium is a constant (grey) or oscillates (with seasonality, green). However, including the behavioural feedback can also lead to periodic waves even in the absence of seasonality. These are triggered by a supercritical Hopf bifurcation (**d**). The frequency of these waves is set by the mitigation delay *τ* (Supplementary Sec. B). Parameters used in **b**,**c**: *β*_0_ = 0.5, *γ* = 0.1. Green: *a* = 0.25, *m*_max_ = 0, *ν* = 1*/*500, Blue: *a* = 0, *m*_max_ = 0.84, *τ* = 30, *ν* = 1*/*100.

**Table 1:** Parameters of the SIRSsm model.

| Parameter | Meaning | Default value | Source |
| --- | --- | --- | --- |
| $\beta_0$ | Basic spreading rate | 0.5 days<sup>-1</sup> | [37] |
| $\gamma$ | Recovery rate | 0.1 days<sup>-1</sup> | [38] |
| $\nu$ | Rate of waning immunity | 0.01 days<sup>-1</sup> | [39] |
| $\tau$ | Mitigation delay | – | Varied |
| $m_{\text{max}}$ | Maximal mitigation | – | Varied |
| $h_{\text{thres}}$ | Threshold hazard | 10<sup>-3</sup> | Chosen |
| $a$ | Seasonal amplitude | – | [26] |
| $\omega$ | Frequency of yearly seasonal variation | $\frac{2\pi}{360}$ days<sup>-1</sup> | – |
| $\epsilon$ | Feedback curvature | 1/4000 | Chosen |

The perceived hazard *h*(*t*) quantifies the risk for oneself and others perceived by people at a given time, providing a proxy for the engagement in mitigation measures; the higher the perceived hazard, the stronger the mitigation. We expect *h*(*t*) to increase with the incidence of the disease, as this sets the risk of getting infected. Explicitly, we calculate *h*(*t*) as the time integral over *I*(*t*), weighted by a delaying kernel *K*_*τ*_ (*t*) with characteristic mitigation delay *τ* :

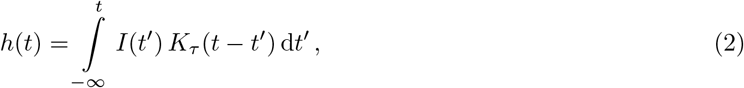

with an Erlang kernel 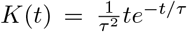 which peaks at *τ* days in the past (Fig. 1a). The mitigation delay *τ* represents both a reaction delay and also a memory time: The reporting of case numbers and the implementation of mitigation strategies does not happen instantaneously. Additionally, individual’s risk perception builds over time and does not only factor in today’s incidence but also that of the previous days and weeks.

We assume that mitigation *m*(*h*) i) increases monotonously with hazard *h*, and ii) saturates at a level *m*_max_ *<* 1 that does not stop transmission completely (i.e., *R*^eff^ *>* 0). A complete, instantaneous stop (*R*^eff^ = 0) would be unrealistic if fractions of the population are agnostic to hazard, and due to logistic limitations. The precise choice of the shape of *m*(*h*) does not impact our results (Supplementary Sec. D).

## Early and late phase effects of mitigation and seasonality

In the classic SIRS model, outbreaks of a disease with *R*_0_ *>* 1 features initially an exponential increase in the incidence *I*. Then, with a decreasing fraction of susceptible individuals, the spread slows down, and eventually the wave of infections is broken. The same is observed in a model including seasonality if the timescale of the outbreak is much shorter than that of the seasons (Fig. 1b). When including behavioural feedback, the dynamics are markedly changed: with increased hazard *h* (and *I*), mitigation becomes stronger, and thereby reduces the spreading rate. Effectively, exponential growth is broken early and the height of the initial wave is reduced (Fig. 1b).

After a transient phase, the incidence is known to stabilise at the stable endemic equilibrium (EE) in the classic SIRS model; seasonality perturbs this equilibrium and induces yearly waves of infections (Fig. 1c, green). This is a classic model result, and also describes well the dynamics of many diseases, like, e.g., the seasonal influenza waves. However, if a disease outbreak is so strong that mitigation becomes necessary, then even without seasonality, oscillations emerge (Fig. 1c, blue). These are triggered by a Hopf bifurcation—purely as a result of the feedback, without accounting for seasonal effects (Fig. 1d) [16].

## Interplay between seasonality and mitigation

In the SIRSm model (without seasonality), the behavioural feedback induces a Hopf bifurcation (periodic waves) if the maximal mitigation *m*_max_ is strong enough to break waves 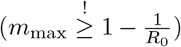 and the mitigation delay *τ* is large enough (Fig. 2a). When including seasonality (SIRSsm model), the Hopf bifurcation curve continuously shifts between the one for winter and the one for summer (Fig. 2b). This leads to a variety of qualitatively different steady states illustrated by four example scenarios. These scenarios represent realisations of the system where mitigation is effectively weaker than, of comparable strength to, or stronger than seasonality.

**Figure 2:**
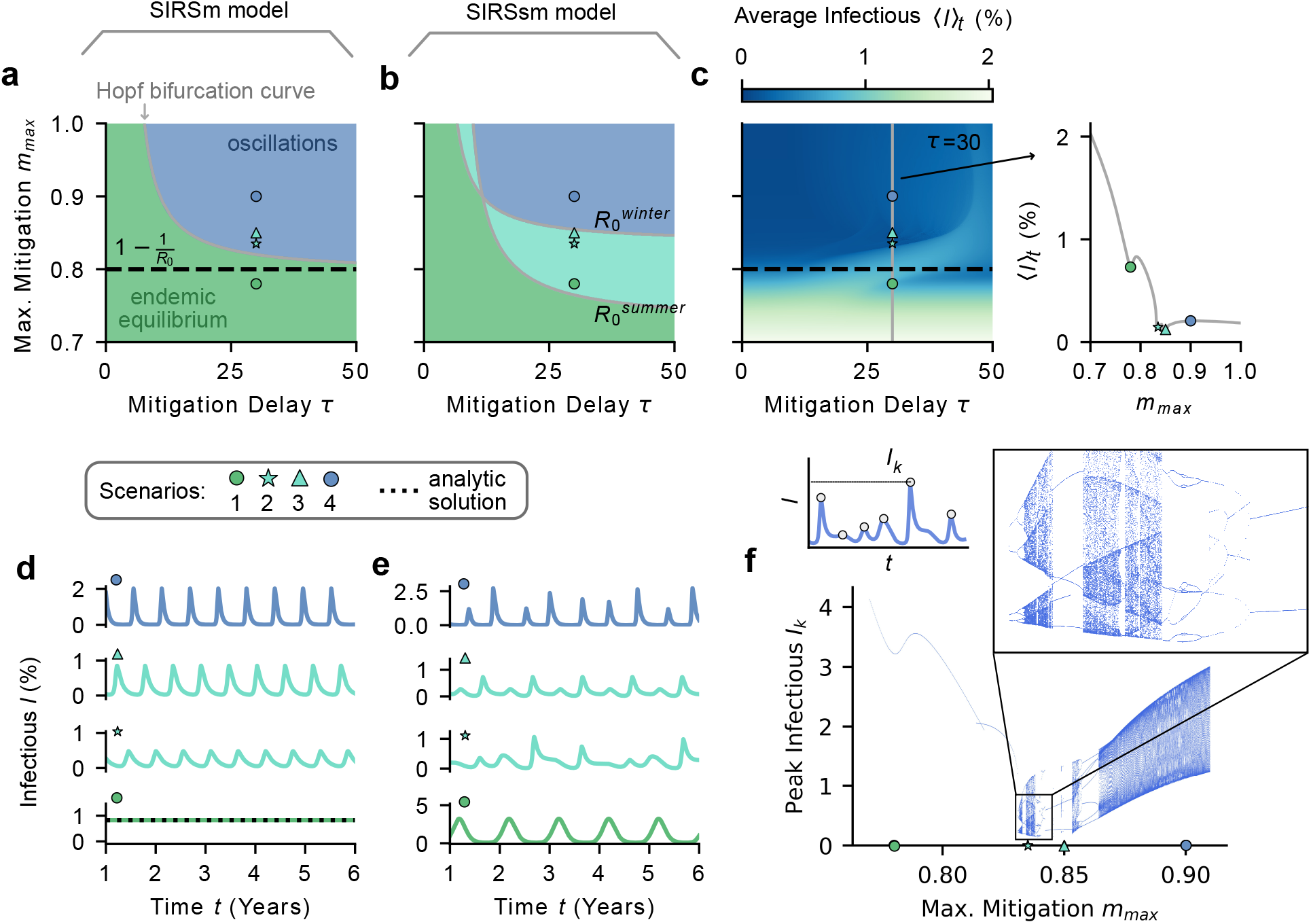
The interplay between seasonality and behavioural feedback opens a rich variety of dynam-ical regimes. We illustrate different regimes in four scenarios indicated by markers. **a)** Without seasonality (i.e., seasonal amplitude *a* = 0), the system has a fixed point that turns unstable through a Hopf bifurcation if maximal mitigation *m*_max_ and delay *τ* are large enough 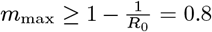 **b)** With seasonality (here *a* = 0.25), the non-autonomous system ceases to possess a fixed point. Computing bifurcation curves with fixed summer- and winter-adjusted spreading rates 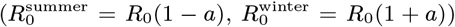 allows to separate different regimes: Seasonality-dominated (green), mitigation-dominated (blue) and balanced (cyan). **c)** Average infections *(I)*_*t*_ (colour coded) in the endemic steady state decrease strongly if mitigation can effectively be stronger than the spread, i.e., if 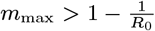 (here computed with seasonality, *a* = 0.25). **d)** In the endemic steady state without seasonality, scenario 1 has a stable equilibrium and scenarios 2-4 display oscillations. **e)** With seasonality, scenarios 1-4 show four qualitatively different dynamics: Sc. 1 shows yearly waves of infections. Sc. 2 shows chaotic dynamics whereas Sc. 3, which only differs slightly in *m*_max_, shows two waves per year. Sc. 4 displays high periodic dynamics. **f)** For a long timeseries, all peak heights are plotted against the parameter *m*_max_, resulting in a peak diagram. Scenario parameters: *a* = 0.25, *τ* = 30, *m*_max_ = 0.78, 0.835, 0.85, 0.9.

If maximal mitigation *m*_max_ is too weak to break a wave (green, Fig. 2a); as a consequence, average infections are high (Fig. 2c). This scenario resembles the classic one without behavioural feedback (Fig. 1c). Without seasonality, the endemic equilibrium is stable and can be approximated analytically by setting *m*(*h*) ≈ *m*_max_ and solving for 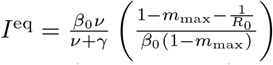 (Fig. 2d, black dotted line). If subject to seasonality, this scenario features yearly waves (green, Fig. 2e).

If mitigation *m*_max_ is sufficiently strong to break waves even in winter 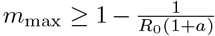 blue in Fig. 2a-f), then the timing and frequency of the waves depend strongly on the mitigation delay *τ* (Supplementary Sec. B). However, seasonality still affects the outbreak sizes, leading to small summer waves and large winter waves. This regime shows periodic waves —potentially of high periodicity— but no chaotic dynamics.

Between the summer- and winter-adjusted Hopf bifurcation curves (cyan, Fig. 2b-f), seasonality and mitigation are of comparable importance. Here, the dynamics is highly sensitive to small parameter changes. As a consequence, qualitatively different dynamics exist close-by in parameter-space, such as phase-locking (two yearly waves in scenario 3) and chaotic dynamics (scenario 2).

One way to display the sensitivity to parameters is to plot the peak heights *I*_*k*_ of a given timeseries against a control parameter, e.g. *m*_max_. By doing so, one obtains a *peak diagram*, similar to a classic orbit diagram that discloses how dynamical regimes change with a model parameter. It reveals that a variation of *m*_max_ around the green and blue scenario hardly change the dynamics, while between the cyan scenarios it can lead to drastically different outcomes, e.g., through period-doubling cascades (Fig.2f). The complex dynamical regimes that go beyond the classic endemic equilibrium, can be understood when viewing the disease model as a driven *epidemiological oscillator*.

## Complex dynamics in the driven epidemiological oscillator

Assume a disease that poses a risk high enough to require strong mitigation. Even in the absence of seasonality, a sufficiently long mitigation delay *τ* then generates self-sustained oscillations, where waves re-surge once mitigation is weakened (Fig. 2d). To this epidemiological oscillator, seasonality adds a periodic driving where the seasonal amplitude *a* represents the coupling strength. Such coupled oscillators are expected to generate complex dynamics [21]. A classical example is the driven Van der Pol (VdP) oscillator [18–20, 22, 23], described by

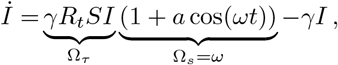

The left hand side of Eq. 3 (if setting *a* = 0) oscillates at its natural frequency Ω_nat_. However, due to the (additive) coupling term on the right, it is stimulated to abide by the external frequency Ω_drive_. Similarly, in the SIRSsm model with mitigation *m* and seasonal amplitude *a*, infections follow

In the absence of seasonality (*a* = 0), the system oscillates at its natural frequency Ω_*τ*_, which is a function of the characteristic mitigation delay *τ* (Sec. B). With seasonality, however, the disease waves are driven (multiplicativey) to synchronise with the seasonal cycle of frequency Ω_*s*_. Hence, the resulting true frequency Ω (if it exists, i.e., if the motion is not quasi-periodic or chaotic) depends on the strength of the seasonal amplitude *a*, which sets the driving strength.

Like in the VdP oscillator, increasing the coupling strength (seasonal amplitude *a*) can lead from high-periodic motion on a torus to chaotic dynamics through period-doubling cascades (Fig. 3a and Supplementary Sec. B). Likewise, Arnold tongues emerge in the SIRSsm model when visualising the averaged number of peaks per year, *W* (the winding number of the system) in the *τ* -*a*-plane. The Arnold tongues manifest as growing regions of phase-locking with increasing *a* (Fig. 3b,c). The shapes of the Arnold tongues depend also on maximal mitigation *m*_max_. If *m*_max_ is strong, the Arnold tongues are well separated, and a cross-sections shows a typical Devil’s staircase (Fig. 3b,d). In contrast, if maximal mitigation is slightly weaker, the Arnold tongues start to overlap and the Devil’s staircase includes abrupt jumps and chaotic regions (Fig. 3c,e). The overlapping of Arnold tongues also gives rise to the coexistence of attractors. This can lead to very different infection dynamics for the same set of parameters (Fig. 3f).

**Figure 3:**
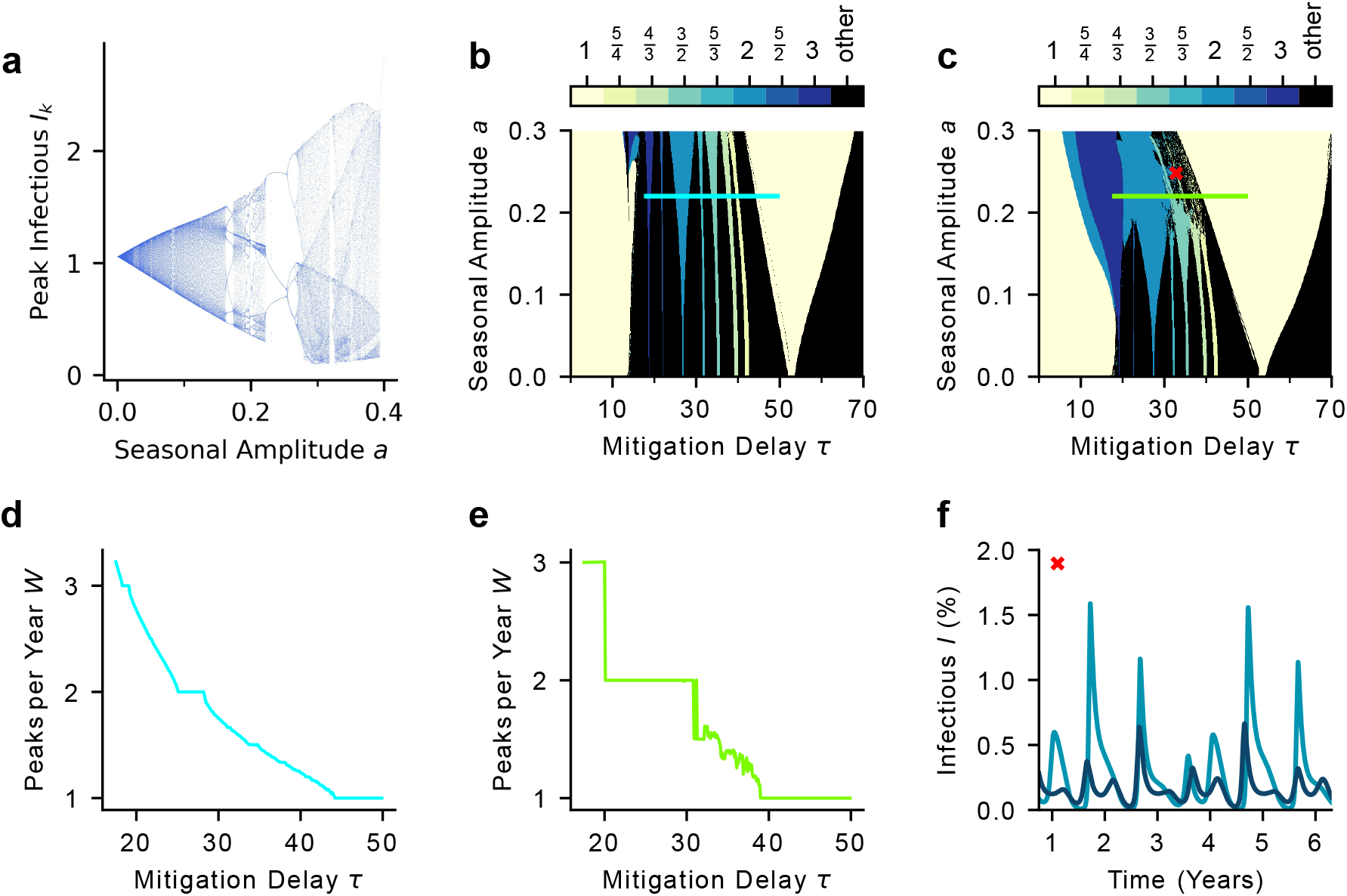
Phase-locking, chaotic wave patterns and coexisting attractors in the SIRSsm model. **a)** A *peak diagram*, i.e., a scatter plot of peak incidences *I*_*k*_ of a long timeseries against the seasonal amplitude *a*. Increasing *a* leads to transitions between high-periodic, low-periodic, and chaotic behaviour. Parameters used: *τ* = 32.5, *m*_max_ = 0.85. **b**,**c)** The averaged number of peaks per year *W* shows Arnold tongues in the *τ* -*a* plane as the result of phase-locking. For strong maximal mitigation the tongues are well separated (*m*_max_ = 0.86, panel **b**) while they overlap for weaker maximal mitigation (*m*_max_ = 0.84, panel **c**). Tongues are displayed only for the ratio of small integers. Cross sections through the Arnold tongues at *a* = 0.22 (green and cyan lines) show a typical Devil’s staircase for *m*_max_ = 0.86 (**d**) but one that includes steep jumps and chaos for *m*_max_ = 0.84 (**e**). **f**) Areas where different tongues overlap give rise to coexisting attractors, i.e., two or more asymptotic states for the same set of model parameters, approached by different initial conditions. The example uses *a* = 0.248, *τ* = 32.8 (red cross in **c**), *S*(0) = 0.521 and *I*(0) = 0.001 vs *I*(0) = 0.361.

Like in the *τ* -*a*-plane, Arnold tongues also emerge in the *τ* -*m*_max_-plane (Fig. 4a). The reason is that decreasing maximal mitigation *m*_max_ has a similar (relative) effect than increasing the seasonal amplitude *a*, as the two are counteracting. For a given disease, we can locate what mitigation strategy a society chooses in the *τ* -*m*_max_-plane: Does it react quickly and resolutely, or slowly and laxly? Of particular interest for disease spread is the regime of sufficiently strong mitigation, where many Arnold tongues with different numbers of peaks per year *W* exist and partially overlap. In this regime, the dynamics is complex and hard to predict as indicated by a number of different measures.

**Figure 4:**
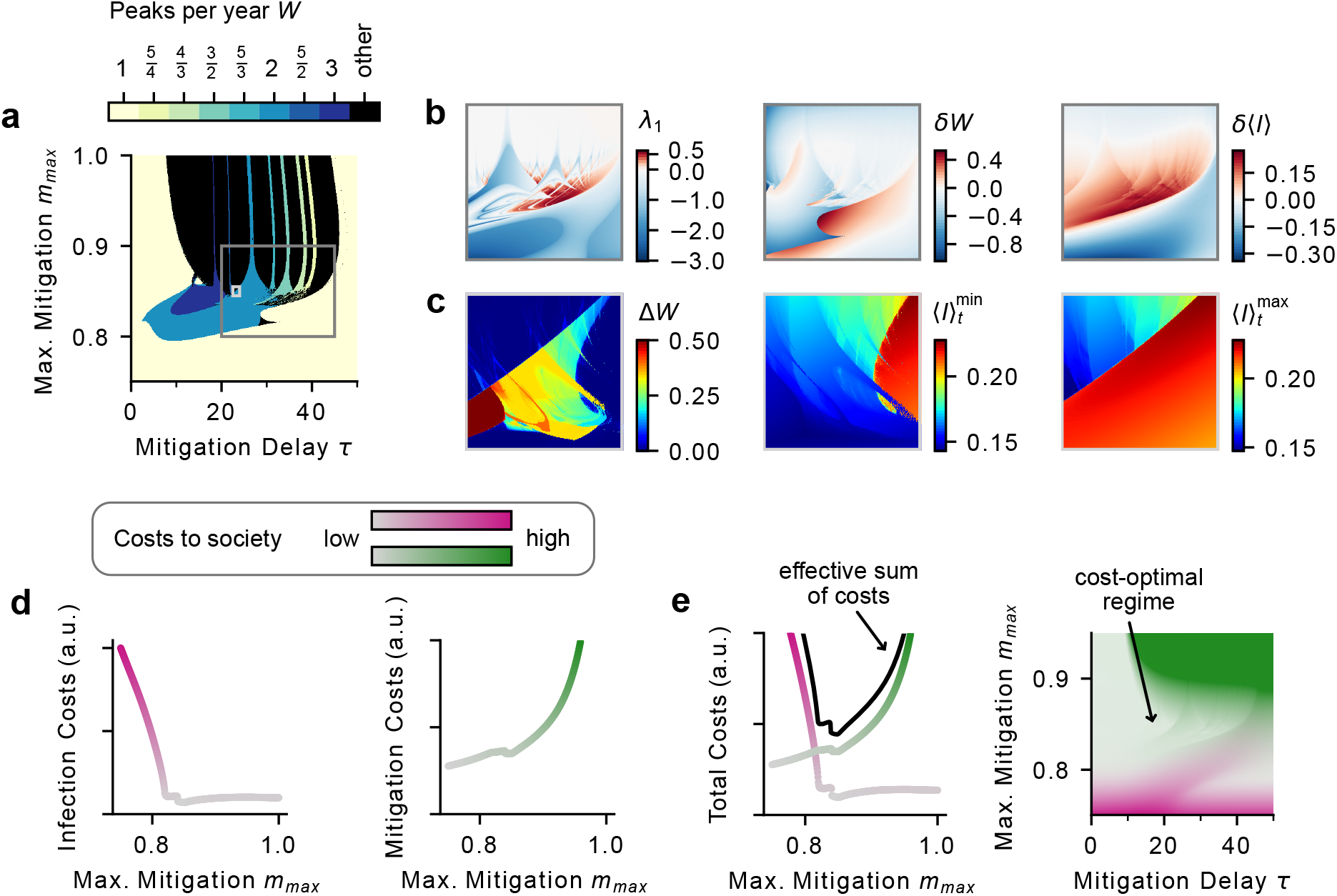
Complex, unpredictable regimes can coincide with cost-optimal regions. **a)** The number of peaks per year *W* in the parameter plane spanned by maximal mitigation *m*_max_ and mitigation delay *τ* shows Arnold tongues. The complexity of the dynamics is characterised by different quantities visualised in two insets (grey rectangles): **b)** Disease prediction is impeded for a large range of parameters (dark grey rectangle in panel a). The largest Lyapunov exponent *λ*_1_ is as large as *λ*_1_ = 0.62 years^*−*1^, which corresponds to a Lyapunov time of 1.6 years (left panel). Parameter variations obtained by Gaussian-weighted samples of trajectories around each point (with widths *σ*_*τ*_ = 4, 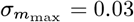) indicate the expected change in peaks per year *δW* as large as *δW* = 0.53 (mid panel) and the expected change of average infections *δ* ⟨*I* ⟩_*t*_ as large as *δ* ⟨*I* ⟩_*t*_ = 0.27% (right panel). **c)** High complexity manifests already in small regions of mitigation parameter space (light grey rectangle in panel a): At fixed parameters both the number of peaks per year *W* and average infections ⟨*I* ⟩_*t*_ differ between coexisting attractors, approached by different initial conditions. This can lead to one additional peak every two years (∆*W* = 0.5, left panel) and up to 50% more infections (local difference between the attractors with minimal infections 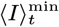 (mid panel) and maximal infections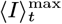 (right panel). **d)** Both infections and the implementation of mitigation strategies come at a cost to society. Assuming that the costs of infections *C*_*I*_ (*t*) are proportional to infections *I*, the average costs over time ⟨*C*_*I*_ ⟩_*t*_ are over-proportionally high if maximal mitigation is not sufficient (here for *τ* = 20). **e** Average costs of mitigation ⟨*C* ⟩)_*t*_ are high for strong maximal mitigation; we assumed costs to diverge for 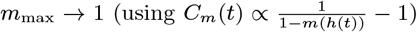 ***f)*** A cost-optimal regime emerges as a balance between the individual costs for infections and mitigation. **g** The cost-optimal region can coincide with the region of chaotic motion and sensitive parameter dependence. The costs are visualised by adapting the colour opacity, not explicitly weighing the costs.

First, chaotic regions arise, where the largest Lyapunov exponent *λ*_1_ is of the order *λ*_1_ ≈ 0.62 years*−*1, which implies a significant divergence given slightly different initial conditions after 1.6 years (Fig. 4b, left panel). Second, any parameter variation —which is to be expected in a realistic scenario— can lead to abrupt jumps between different dynamical regimes. Gaussian noise around a fixed parameter value with variances *σ*_*τ*_ = 4 and *σ*_*m*_max=0.03 can lead to expected changes of the number of peaks per year as high as *δW* = ±0.5, and expected increases in average infections as high as *δ*(*I*)_*t*_ = 0.27%, compared to a fixed parameter value (Fig. 4b, mid and right panels). Third, even within small regions of parameter space the coexistence of attractors provides another reason for uncertainty. Depending on initial conditions, the number of peaks per year can differ by as much as one peak every two years (∆*W* = 0.5, Fig. 4c, left), and the number of average infections can differ by up to 50% (Fig. 4c, middle and left). Even when following the ‘optimal’ attractor, the minimal average infections can change by about 50% for tiny changes in parameters (Fig. 4c, middle) - similar for the ‘worst’ attractor (Fig. 4c, right). While complexity arises only in some portion of parameter space, we will show next that it might well be a region of particular importance.

## Overlap of cost-optimal and complex regimes

Besides the mathematical existence of complex disease states, there is another dimension to the question: What kind of mitigation strategy employed by a society is to be expected, i.e., which areas of parameter space are epidemiologically relevant? Intuitively, mitigation must be adequate to the disease, i.e., it must be the cost-optimal strategy from societal viewpoint. In this section, we show that this cost-optimal strategy may lie right in the region of complex and unpredictable disease dynamics, if accounting for costs associated to infections as well as mitigation.

A precise quantification of the multifaceted costs of infections and mitigation to society is practically impossible. We therefore take a first-order approximation: We first assume that every single infection incurs the same expected cost, which represents all potential consequences of the infection, from sick-leave to hospitalisation, death, and long-term health implications. Thus total costs of infections *C*_*I*_ (*t*) are proportional to the total number of infections, i.e., *C*_*I*_ (*t*) ∝ *I*(*t*).

The time-averaged infection costs (*C*_*I*_ (*t*))_*t*_ are particularly high if mitigation is not sufficiently strong to break waves on its own, namely if *m*_max_ 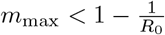 (Fig. 4d). Once maximal mitigation is strong enough, the costs remain low in a wide regime (Fig. 4d). However, implementing resolute mitigation strategies is costly for the economy, education, culture, and individual well-being [4]. We assume that little mitigation is easily achievable, whereas stronger mitigation becomes increasingly more costly, e.g., because schools would need to be closed or contacts would need to be strongly reduced. Thus we define mitigation costs *C*_*m*_(*t*) to be zero when the disease is not mitigated at all, and diverging if it would be completely mitigated instantaneously.

Infection costs drop steeply once maximal mitigation *m*_max_ is strong enough, and thus the cost-optimal region for society is located where disease mitigation is “just enough” to reach the flat part of the infection costs (Fig. 4f,g). This part overlaps with the region of complex dynamics determined previously. Here, chaotic dynamics, high sensitivity to parameter changes, and the coexistence of attractors strongly affect disease forecasts. Furthermore, already slight deviations from the optimal regime can significantly increase the average number of infections (Fig. 4f). For example, a change of only about ∆*m*_max_ = 0.05 from scenario 1 to 2 (Fig. 2d) increases the average number of infections almost five-fold. In other words, each individual is, on average, infected almost five times more often.

The definitions of the costs we used omit nonlinear effects in disease burden due to hospital overload, and hysteresis effects in mitigation, i.e. prolonged resolute measures becoming more costly with time. Nevertheless, due to the pronounced increase of infections at insufficient maximal mitigation, we argue that the location of the cost-optimal regime close to the sudden increase in average infections is very stable against the definition of the cost functions.

### Signatures of complex dynamics in influenza and COVID-19

Our theoretical results above indicate that strategies minimising the total costs of infections and mitigation can drive societies to a regime of high parameter sensitivity and chaos. But, can we find signatures of that in real world data? To investigate this, we analysed the wave patterns of COVID-19 and seasonal influenza (Supplementary Sec. C) to compare them with those obtained from our model.

Influenza is a disease with marked seasonality but, compared to COVID-19, with little mitigation measures implemented by society. Analysing European influenza data, we observe generally one (seasonality-driven) peak per year, typically between January and March (Fig. 5a,c, grey). In contrast, COVID-19 waves (in the pre-Omicron era) required active mitigation of a strength comparable to the seasonal variations [24–26]. Here, we found a broad distribution of wave frequencies, i.e. the inverse of the time between consecutive waves, as well as of the waves’ respective timings within the year, indicative for complex disease dynamics (Fig. 5b, d, grey). This is in stark contrast to influenza and the classic endemic state, and is qualitatively in line with our theoretical results that one observes complex, potentially chaotic wave patterns when seasonality and mitigation have a comparable impact.

**Figure 5:**
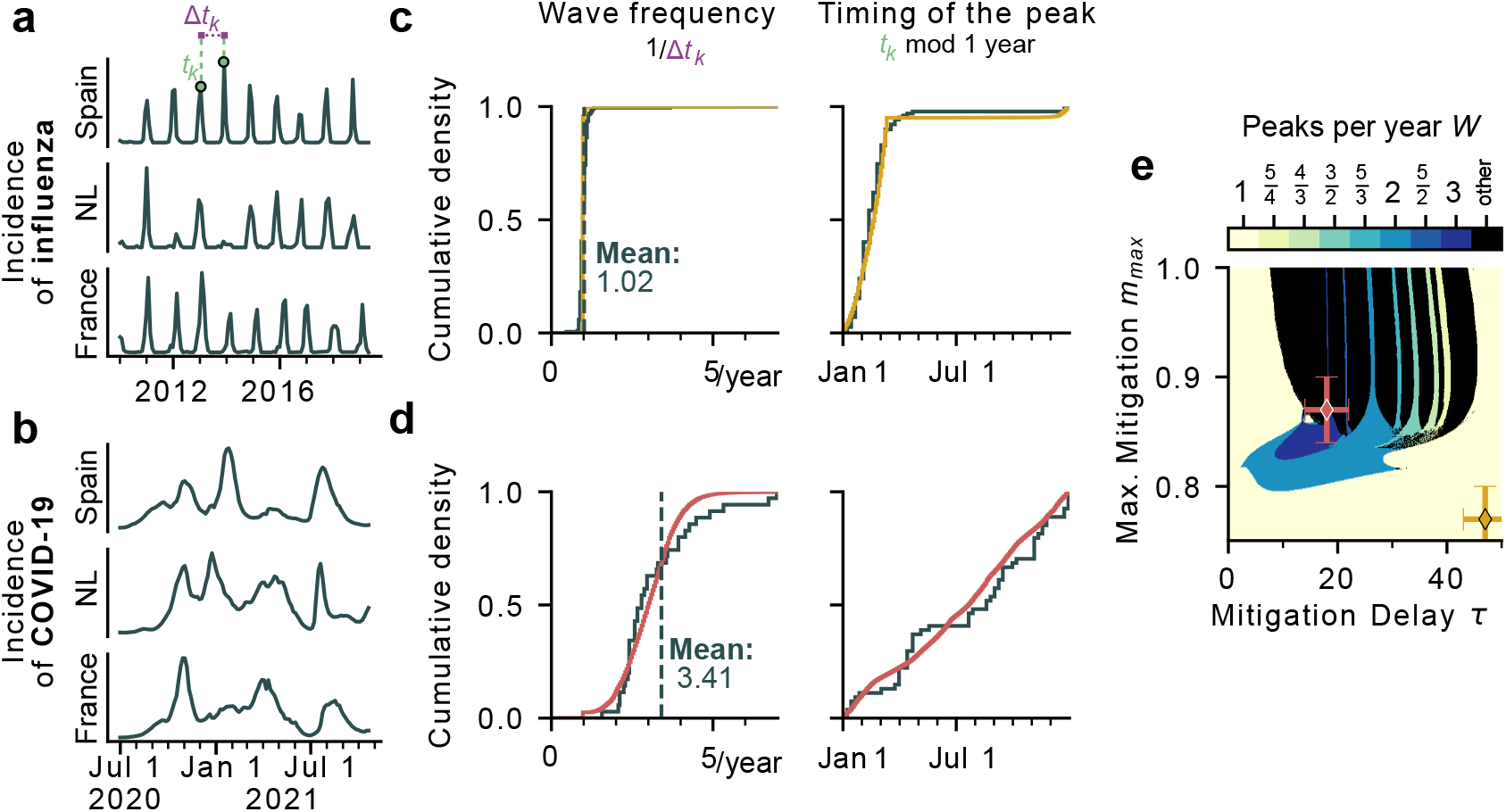
Wave characteristics in northern countries suggest that the spread of COVID-19 lied within the regime of complex dynamics. **a**,**b)** We collected timeseries of COVID-19 (summer 2020 until the spread of the Omicron variant) and seasonal influenza (2010 to 2019) incidence in several northern countries (Supplementary Sec. C) and extracted the wave frequency, i.e., the inverse time between consecutive waves, and the time of the year at which incidences peaked. **c)** Influenza waves show a narrowly peaked distribution of wave frequencies around 1 peak per year with this peak occurring predominantly between January and March. **d)** Wave frequencies show a broad distribution from 1 to 7 peaks per year in COVID-19 data. These peaks were observed all year long. **e)** For comparison with our model, we sampled trajectories around points in the *τ* -*m*_max_-plane with Gaussian weights to also extract the wave frequencies and peak timings in simulations. The resulting model distributions resemble those observed for COVID-19 in the regimes of complex dynamics (red in d,e) and those of influenza in the regime of yearly peaks and weak mitigation (yellow in c,e). Errorbars in e) indicate the widths of the Gaussian sample (*σ*_*τ*_ = 4, 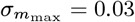.

When comparing the experimental data quantitatively with our model, we first have to acknowledge that the spread of influenza and COVID-19 did not occur in a “stationary” society, akin to a model with a fixed parameter set. To reflect such parameter variability in the model, we chose to obtain the model characteristics (wave frequency and timing) from a Gaussian-weighted sample of trajectories (with widths of *σ*_*τ*_ = 4 and 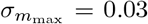 in the *τ* -*m*_max_-plane (Fig. 5e). It shows influenza-like dynamics (a single yearly infection wave between January and March) when seasonality is stronger than mitigation (small *m*_max_, Fig. 5c,e yellow). Importantly, broad distributions of the number of yearly waves and their respective timing, as for

COVID-19, are best reflected by the complex regime of our model where diverse dynamical regimes co-exist (moderate *m*_max_, Fig. 5d,e red). While this is comparing the pandemic (initial) phase of COVID-19 with the endemic steady-state in our model, a comparison to the initial transient phase of the model yields very similar results (Supplementary Fig. 7). Further, data of the Omicron variant shows qualitatively similar distributions (Supplementary Fig. 7). Overall these results suggest that the dynamics of COVID-19 might indeed have played out in a sensitive and highly complex region of (real-world) parameter space and that for diseases where mitigation of outbreaks remains necessary for years, irregular and complex dynamics might result even in their endemic phase.

### A general mechanism to complex endemic states

The emergence of complex dynamics in endemic diseases has been reported before. For example, a great body of literature has focused on explaining characteristic wave patterns of diseases such as measles or chickenpox that often show yearly outbreaks of alternating peak heights or biennial wave patterns [27–31]. In particular, the debate on whether such observations are the result of chaotic dynamics or just stochasticity [32, 33] has led to several works demonstrating the emergence of period-doubling cascades to chaos in models with seasonality, when increasing the seasonal amplitude [27, 29–31]. However, the emergence of complex dynamics in these models typically relies on high reproduction numbers and does not account for disease mitigation as the result of human behaviour. We explain the emergence of complex dynamics by the interplay of two essential parts of disease spread, providing a mechanism guaranteed to lead to complex behaviour when seasonality and disease mitigation are of comparable strength, which is also applicable for lower spreading rates (e.g., *R*_0_ = 2, Supplementary Sec. F).

Performing a state- and parameter space analysis with any dynamical system becomes vastly more difficult with increasing complexity of the model. Thus, we decided to neglect effects that we considered inessential for the purpose of our study. For instance, we decided not to include vaccination explicitly; assuming regular or seasonal vaccination, one can instead see vaccinations as influencing the spreading rate or the amplitude of seasonality, respectively. Further, evolving viral variants, which would further drive unpredictability, are only partially reflected in the relatively high rate of waning immunity *ν* = 1*/*100 days^*−*1^. Arguably the largest simplification in our model however, is the behavioural feedback condensing the highly erratic component of human behaviour into just a few simple functions: The hazard *h* as a weighted delay of past infections, and the shape of mitigation *m*(*h*) in response. For instance, we do not distinguish between different types of disease mitigation such as voluntary contact reduction vs official non-pharmaceutical interventions. This is only approximately captured by the weighted delay in the feedback. We also assumed that disease mitigation and seasonality act independent of each other, which allows to write them as multiplicative factors on the spreading rate. This assumption excludes the possibility of explicit seasonal awareness, namely preemptive mitigation at the onset of winter, which is only implicitly accounted for by the value of the seasonal amplitude *a*. Similarly, the delayed feedback mechanism excludes the anticipation of future events, namely adapting behaviour in light of expecting increasing incidences (Supplementary Sec. E). The precise functional shape of the mitigation *m*(*h*) however, only impacts the dynamics quantitatively. If mitigation measures are implemented at low infection numbers and are strong enough, complex dynamics emerge for a variety of different functional shapes *m*(*h*) (Supplementary Sec. D). We thus argue, that our qualitative results are general and independent of the precise feedback mechanism: Human behaviour can be the cause of highly complex endemic disease states, as the result of balancing costs of infection and mitigation.

## Data Availability

All code to reproduce the analysis and figures shown in the manuscript as well as in the supplementary information will be available online on GitHub https://github.com/Priesemann-Group/chaosproject

## Acknowledgements

All authors received support from the Max-Planck-Society. SC, JW, SB and VP acknowledge funding from BMBF, RESPINOW (Project 031L0298) and infoXpand (Project 031L0300A) consortia.

## A Differential equations

The modified SIRS model (SIRSsm model) used for the analysis throughout this manuscript is described by a set of deterministic differential equations. Its solution is unique and relies solely on the initial conditions. Following a common approach [13, 34, 35], the hazard *h* used in the behavioural feedback is given by the convolution of the infectious compartment *I* with a delaying kernel *K*:

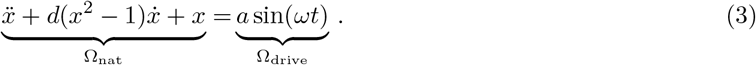

By using an Erlang kernel of second order that peaks at *τ* days in the past 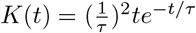 one can reduce the set of integro-differential equations for *S, I* and *R* to a set of ordinary differential equations [36] for *S, I, R, h*^*’*^ and *h*, where *h*^*’*^ is an auxiliary compartment. Hence, the time evolution of the hazard *h* does not require the calculation of the convolution integral, which allows to solve the differential equations numerically considerably faster.

Mitigation *m*(*h*) is given by a softplus function that increases linearly for *h < h*_thres_ and saturates thereafter [10]:

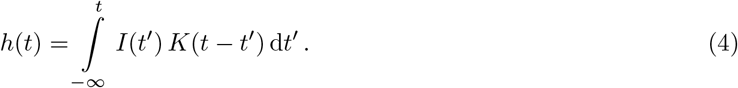

*h*_thres_ is the threshold hazard beyond which no further increases in mitigation are possible. The third parameter defining *m*(*h*) is *E*, which defines the smoothness of the transition from linear increase to constant value.

Additional to the four parameters defining the feedback (*τ, m*_max_, *h*_thres_, *E*) and the two defining the seasonal forcing (*s, ω*), the dynamics requires the specification of three transition rates. Those are given by the basic spreading rate *β*_0_, the recovery rate *γ* and the rate of waning immunity *ν*. The rates *β*_0_ = 0.5 and *γ* = 0.1 were set such that the resulting basic reproduction number equals *R*_0_ = 5, which lies in the range of R-values estimated for SARS-CoV-2 [37, 38]. The waning rate *ω* = 1*/*100 was set such that on average, an individual is immune for 100 days, incorporating the emergence of new variants which can bypass existing immunity, and thus effectively reduce the period of immunity.

Given a set of initial conditions, the dynamics is thus fully described by

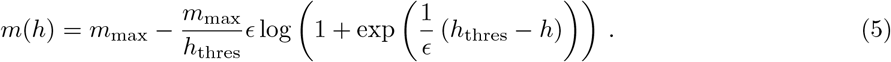

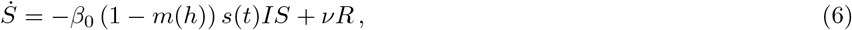

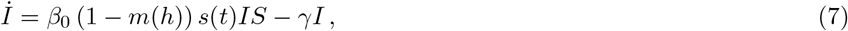

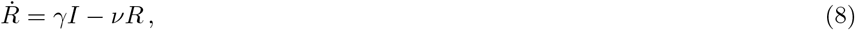

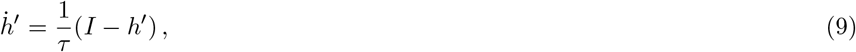

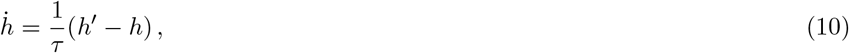

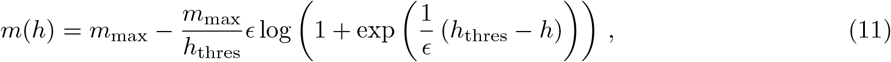

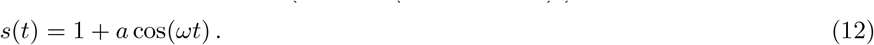

For most of the analysis performed throughout this manuscript, the initial conditions were set to *I*(0) = *h*^*’*^(0) = *h*(0) = *h*_thres_, *S*(0) = 1 − *h*_thres_ and *R*(0) = 0. More details are provided in Sec. B. A full collection of all parameters and compartments in the model is given in Tab. 1 and Tab. 2, respectively.

**Table 2:** Model compartments and functions.

| Variable | Meaning | Definition |
| --- | --- | --- |
| $S$ | Fraction of susceptible individuals | – |
| $I$ | Fraction of infectious individuals | – |
| $R$ | Fraction of recovered individuals | – |
| $\beta$ | Effective spreading rate | $\beta(h) = \beta_0 (1 - m(h))$ |
| $h$ | Perceived hazard by society | $h(t) = \int_{-\infty}^t I(t') K(t - t') dt'$ |
| $K$ | Delaying kernel | $K(t) = \frac{1}{\tau^2} t e^{-t/\tau}$ |
| $m$ | Mitigation due to human behaviour | $m = m_{\text{max}} - \frac{m_{\text{max}}}{h_{\text{thres}}} \epsilon \log \left( 1 + \exp \left( \frac{1}{\epsilon} (h_{\text{thres}} - h) \right) \right)$ |
| $s$ | Seasonality | $s(t) = 1 + a \cos(\omega t)$ |
| $X_k$ | Quantity $X$ at wave peak $k$ | $X$ at the $k$th occurrence of $\dot{I} = 0 \wedge \ddot{I} < 0$ |
| $T$ | Effective period time of the oscillation | $T = \lim_{n \rightarrow \infty} \frac{1}{n-1} \sum_{k=1}^n (t_{k+1} - t_k)$ |
| $\Omega$ | Effective frequency of the oscillation | $\Omega = \frac{2\pi}{T}$ |
| $W$ | Peaks per year or winding number | $W = \frac{\Omega}{\omega}$ |

## B Dynamical regimes

### Stability of the endemic equilibrium

The endemic equilibrium of the system is the fixed point EE = { *S*^*∗*^, *I*^*∗*^, *R*^*∗*^, *h*^*’∗*^, *h*^*∗*^ } defined by 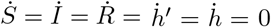 . In order to study the stability of the endemic equilibrium, the seasonal forcing *s*(*t*) needs to be set to *s* ≡ 1 (no seasonal amplitude, *a* = 0), otherwise the system does not possess a fixed point. The endemic equilibrium is then numerically computable via

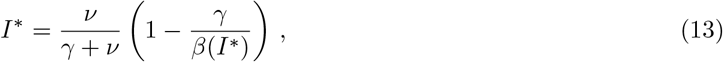

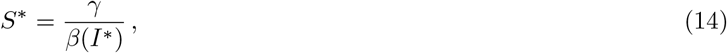

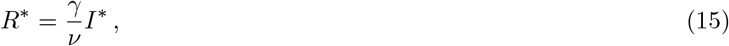

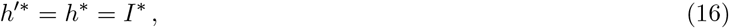

where we wrote β(I∗)= β0(1−m(I∗)).

Its stability can be computed by evaluating the Jacobian *J* of the system at the endemic equilibrium:

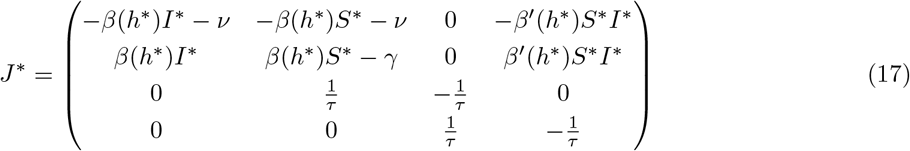

If there is an eigenvalue with positive real part, the endemic equilibrium is unstable; if not, it is stable. In the case of no delay, namely *h* = *I*, the endemic equilibrium is always stable, because *m*^*’*^(*I*) *>* 0 and hence *β*^*’*^(*I*) ≤ 0 [40].

### Peaks per year *W*, resemblance to VdP oscillator

Among other measures, we chose the averaged number of peaks per year *W* as a measure to distinguish different dynamical regimes. It is calculated by initialising the system, waiting until the system has settled into its asymptotic state and collecting *N* peaks *I*_1_, …, *I*_*N*_ at times *t*_1_, …, *t*_*N*_ . The averaged number of peaks per year is then given by *W* = 360 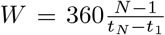 Note that if the motion is periodic (i.e., not chaotic), this is equivalent to the notion of a winding number in the driven Van der Pol oscillator 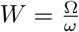 [18] with Ω the frequency of the system and 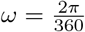

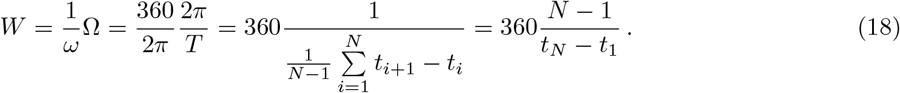

Without seasonality, the period of the Hopf oscillations increases linearly with mitigation delay *τ* (Fig. 6b). In contrast, maximal mitigation *m*_max_ has less influence on the period *T* (Fig. 6c). At the bifurcation point, the later is given by the imaginary part of the bifurcating eigenvalues of the Jacobian. However, this approximation quickly deteriorates when moving away from the bifurcation curve (Fig. 6a-c).

**Figure 6:**
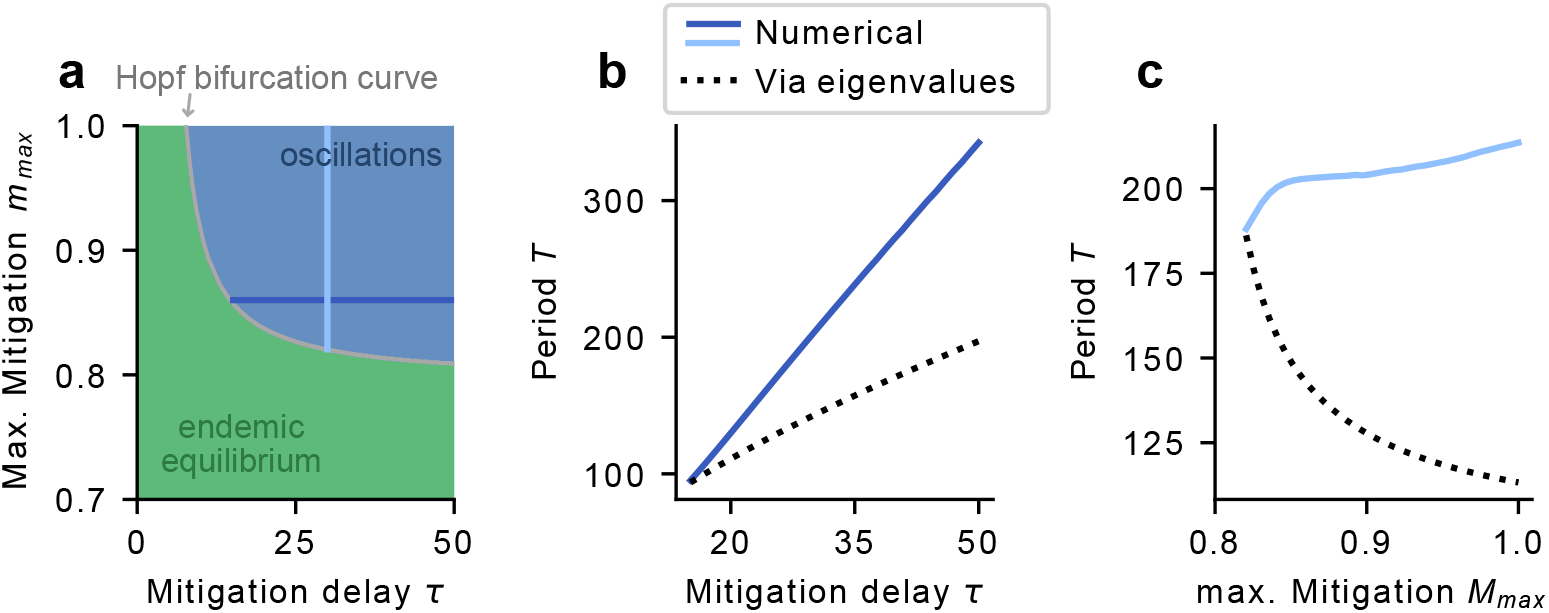
The period of the SIRSm model (no seasonality) increases linearly with the mitigation delay. **a)** For large mitigation delay *τ* and strong maximal mitigation *m*_max_ the endemic equilibrium turns unstable through a Hopf bifurcation. The period of the oscillation at the bifurcation point is given by the imaginary part of the bifurcating eigenvalues of the Jacobian. **b)** The period of the oscillations increases linearly with the mitigation delay *τ*, though the approximation through the eigenvalues quickly deteriorates beyond the bifurcation curve. **c)** Maximal mitigation *m*_max_ does not influence the period of the oscillations as much.

While there exists a clear resemblance between the driven VdP oscillator and our model, the systems differ in several aspects. In the driven VdP oscillator the external frequency *ω* can be adjusted and be used as a control parameter, namely Arnold tongues emerge in the *ω*-*a* plane where *a* is the coupling strength. Contrarily, in the SIRSsm model the external frequency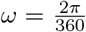describing the yearly seasonal change is fixed, but the internal frequency can be changed by varying *τ* . Additionally, the coupling acts additively in the driven VdP oscillator but multiplicatively in the SIRSsm model. Despite these differences however, the two models show very similar behaviour. For instance, if the coupling strength, i.e., seasonal amplitude is weak in the SIRSsm model, (high-)periodic motion dominates. Here, seasonality influences the peak heights induced by the behavioural feedback, but does not enforce yearly waves on its own. The dynamics is bound to an invariant torus in state space, which leads to (high-)periodic (i.e. *p* waves in *q* years) or quasi-periodic wave patterns, depending on the ratio between the mitigation-induced frequency Ω_*τ*_ and the seasonal frequency *ω*.

### Largest Lyapunov Exponent

The dynamics of a dynamical system such as the SIRSsm model is chaotic when the largest Lyapunov exponent *λ*_1_ of that system is larger than zero. We compute it by evolving a system state 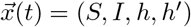 and a perturbation vector 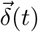 in parallel [20]. The system state is evolved with the set of equations of the SIRSsm model (Eq. 6-10) whereas the perturbation is evolved according to the linearised set of equations:

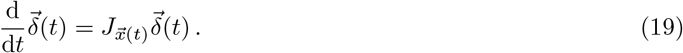

Here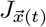 is the Jacobian of the system at the state 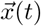 given by

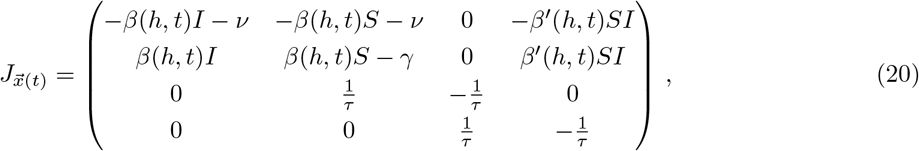

where 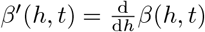 In practice, the set of equations Eq. 6-10 is evolved for 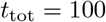 years such that the trajectory has settled onto the attractor. Subsequently, the set of equations is extended by the four equations of Eq. 19 to integrate the state and the perturbation in parallel. Using 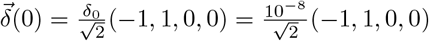 the extended set of equations is integrated for another 100 years such that the perturbation vector converges to the direction of fastest growth. After rescaling of the perturbation to its initial size *δ*_0_ = 10^*−*8^, the calculation of the Lyapunov exponent is performed. For this purpose, the extended system is evolved *n* = 20 times for *t*_tot_ = 100 years. After each integration, the perturbation vector is rescaled to its initial size and the largest Lyapunov exponent is calculated by 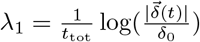

Subsequently, the *n* = 20 results are averaged to obtain an accurate measurement of the largest Lyapunov exponent of the attractor.

### Coexisting attractors

Depending on the initial conditions, the dynamics of the SIRSsm model can settle into different asymptotic states. Those can differ in one or multiple characteristics of the asymptotic dynamics, e.g., by a different number of peaks per year *W* or average infections ⟨*I* ⟩_*t*_ (Fig. 4).

In order to find coexisting attractors, we solved the differential equations of the SIRSsm model for a two-dimensional grid of different initial conditions. In practice, we chose *S*(0) and *I*(0) in the domains *S*(0) ∈ (0, 1) and *I*(0) ∈ (0, *S* (0)) for a total of 100 initial conditions. The other initial conditions were set to *R*(0) = 1 − *S*(0) − *I*(0) and *h*^*’*^(0) = *h*(0) = 0. Even though this method does not scan the whole state space for the existence of coexisting attractors, the choices of initial conditions represent a grid of all physical starting conditions, without any hazard at the outbreak of the disease. Note that the exact number of coexisting attactors can not be estimated using this method, but it is sufficient to reveal the existence of at least two coexisting attractors by quantifying differences in the dynamics such as *W* or ⟨*I* ⟩_*t*_. ⟨*I* ⟩_min_ and ⟨*I* ⟩_max_ in Fig. 4 refer to the minimal and maximal average number of infections found between coexisting attractors for a single set of parameters. ∆*W* is the difference between the minimal and maximal number of peaks per year between coexsiting attractors.

### Magnitude of uncertainty regarding disease prediction

Aside from the many simplifying model assumptions made for the SIRSsm model, real-world disease dynamics is undoubtedly not characterised by one specific parameter set of the SIRSsm model. Rather, the way a society mitigates a disease fluctuates and changes over time, implying that a single timeseries using one specific parameter set is not representative. Therefore, when comparing model simulations to data or to quantify parameter sensitivity, we chose to obtain wave characteristics and expected changes of the dynamics by considering parameter fluctuations around each point in the *τ* -*m*_max_-plane. In practice, we calculated the expected change in the number of peaks per year *δW* and in the number of average infections *δ* ⟨*I* ⟩ between Gaussian-weighted samples of trajectories around each parameter combination. We assumed fluctuations to be Gaussian with standard deviations of *σ*_*τ*_ = 4 and 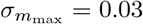 Numerically, this was achieved by first computing the number of peaks per year *W* and number of average infections (*I*) in continuous resolution (Fig. 2c, Fig.4a) and applying a Gaussian filter to it (Fig.4b).

## C Comparison to influenza and COVID-19 data

As our model does not attempt to reproduce exact timeseries of real disease dynamics, we considered ensemble characteristics of waves of infections to compare the SIRSsm model to data. Thus, we considered the *wave frequency*, i.e. the inverse of the time between consecutive infection waves, and the timing of the waves during the year. We defined the time of a wave as the time of its peak. Note that although the wave frequency has the same units as the number of peaks per year *W*, there is a subtle difference: *W* is the number of peaks per year *averaged* over a timeseries, while the wave frequency is the inverse time between a peak and the next, *defined for each peak* in a given timeseries. In the case of 2 peaks per year for instance, *W* and the wave frequency only agree when the two peaks occur in regular half-year intervals.

To obtain cumulative distributions for COVID-19 (Fig. 5) we took the daily new confirmed cases (smoothed by 7 day rolling average) from ourworldindata [41] for 18 northern countries (Tab. 3). We only considered the waves from summer 2020 (July 1, 2020) up to the first waves caused by the Omicron variant (with the cut chosen to be October 20, 2021). This way, we left out the first waves in spring 2020, since they marked the initial outbreak, as well as waves caused by Omicron, since it marked a considerable change in both disease dynamics and mitigation efforts not covered by our model. The period when Omicron dominated displayed different but qualitatively similar characteristics (Fig. 7). Ideally, we would have cut out the phase of mass-vaccinations in most of the considered countries in 2021 as well. However, this would have presented a drastic reduction in the size of our dataset. We argue that even in the first months after large-scale immunisation behavioural changes adapted slowly and many mitigation efforts remained.

**Table 3:** Data sources for Influenza and COVID-19 timeseries.

|  | Timeframe | Countries | Source |
| --- | --- | --- | --- |
| COVID-19 | Summer 2020 (July 1, 2020) to before the first Omicron waves (October 20, 2021) | Italy, Spain, Portugal, Israel, Germany, France, USA, Canada, Switzerland, Austria, Netherlands, Belgium, Poland, Czechia, Croatia, Denmark, Sweden and Lithuania (18 in total) | [41] |
| Seasonal influenza | 2010–2019 | Austria, Belgium, Bulgaria, Czechia, Germany, Denmark, Estonia, Greece, Spain, France, Hungary, Ireland, Lithuania, Latvia, Netherlands, Norway, Poland, Portugal, Romania, Sweden, Slovenia, Slovakia, Italy, Finland (24 in total) | [7] |

**Figure 7:**
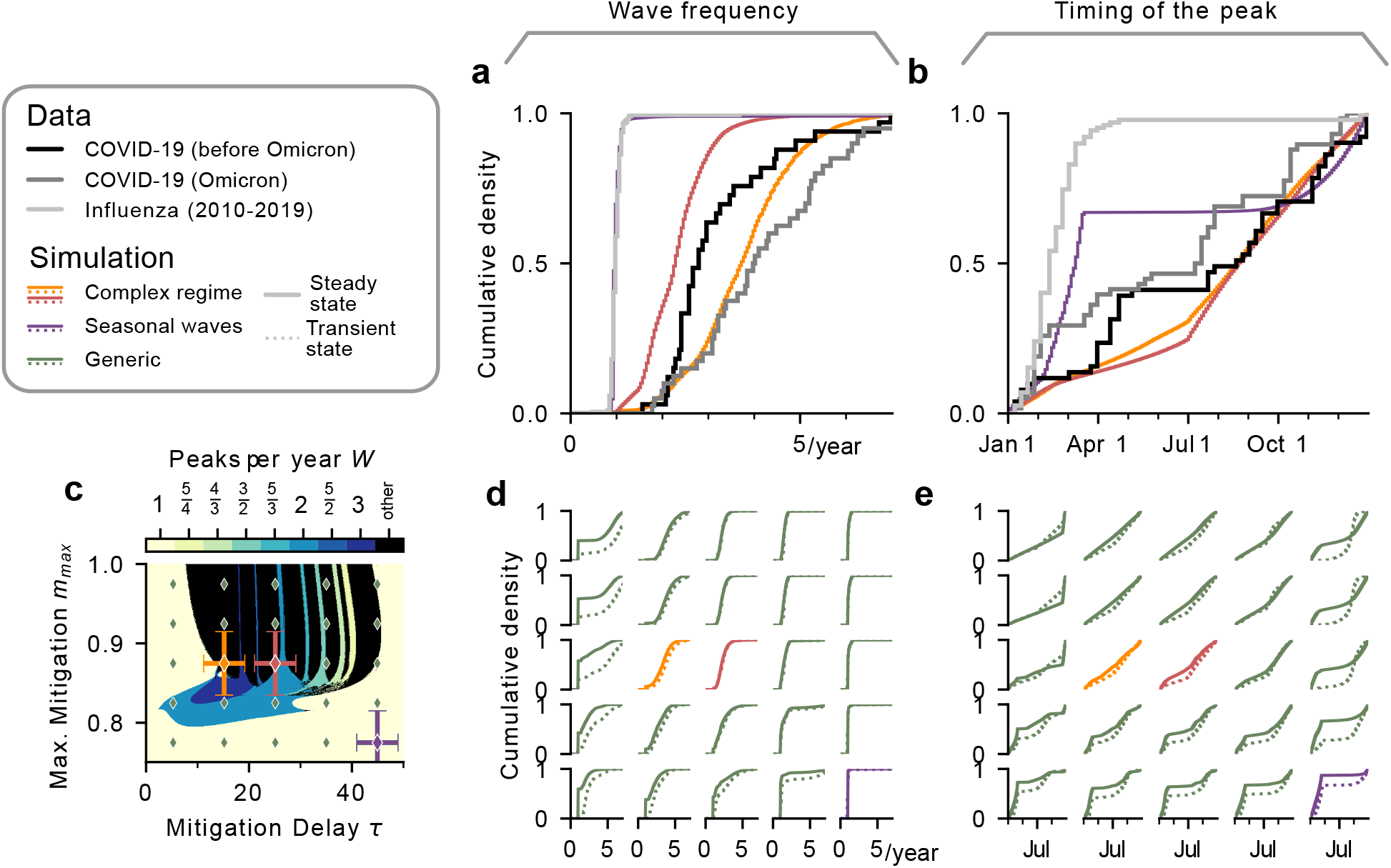
Wave characteristics quickly equilibrate to their steady-state distributions. In Fig. 5 we compared data from the first two years of COVID-19 (leaving out the very first outbreaks and stopping when the Omicron variant emerged) to steady-state model simulations. Here we check the validity of this approach by simulating the model only for two years (leaving out only the first 180 days, i.e. the first outbreaks) and also compare to data for the Omicron variant. Full, coloured lines represent the steady-state distributions as in Fig. 5, dashed lines the two year simulations. **a,b)** COVID-19 spread for the Omicron variant (grey curve) has a slightly different distribution of wave frequencies and wave timings than for pre-Omicron COVID-19 (black curve), but is still close to distributions of model simulations in the complex regime (orange and red curves). **c-e)** For both the wave frequencies and the wave timings the model parameter sets that match COVID-19 data before Omicron qualitatively in the steady state also match the data in the two-year simulations and for Omicron. A 5×5 grid is placed onto the *τ* -*m*_max_-plane and distributions are sampled around the centre points of the grid. The distributions are only slightly affected by the steady-state assumptions all over parameter space.

We obtained the wave peaks via the *find_peaks_cwt* method in the *scipy*.*signal* package of Python 3.7. with 25-50 days set as the expected width of the peaks of interest. Since the algorithm sometimes returned slightly shifted peaks compared to a manual bare-eye check, we took the returned peak positions and corrected them by extracting the position of the maximum of the 40 data points in the immediate vicinity. Furthermore, we considered two peaks within 1 month as being the same up to infection and reporting noise in the data. The distributions of wave characteristics were in the end obtained from an ensemble average of all countries considered.

To obtain cumulative distributions for seasonal influenza (Fig. 5), we took sentinel detection timeseries from 2010 to 2019 in 24 European countries (Tab. 3) with weekly resolution from the Surveillance Atlas of Infectious Diseases of the European Centre for Disease Prevention and Control (ECDC) [7]. This way, we filtered out the earlier years with lacking data and years beginning from 2020, in which the spread of Influenza was significantly affected by the mitigation efforts to contain COVID-19. We obtained the peaks using the same procedure as for COVID-19 with 5-6 weeks set as the expected width of the peaks of interest, corrections of the peaks within the 6 data points in the immediate vicinity, and considered two peaks within 10 weeks to be the same up to noise.

In the simulations, we took ensembles of trajectories in the *τ* -*m*_max_-plane, centred around different base values in this two dimensional parameter space. For each trajectory we computed the wave frequencies and timing of the waves in the steady state. We computed the cumulative distributions, weighing the trajectories with a multivariate Gaussian distribution with standard deviations of *σ*_*τ*_ = 4 and *σ*_*m*_max = 0.03. In principle, one should rather compare the initial, transient phase of the model simulations to the data of COVID-19. However, the model trajectories very quickly converge to the steady-state characteristics (Fig. 7).

## D Different feedbacks

### Logistic feedback

The softplus feedback *m*(*h*) (Eq. 5) used for the analysis throughout this manuscript increases linearly for small *h* as soon as *h >* 0. One could argue that a certain threshold hazard should be required in order to trigger mitigation. A suitable feedback reflecting this notion could be given by a logistic feedback of the three known parameters *m*_max_, *h*_thres_ and *E*:

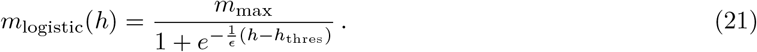

Using this different definition, oscillations emerge again for strong maximal mitigation *m*_max_ as well as long mitigation delay *τ* (Fig. 8a,b). However, the Hopf bifurcation curve is shifted to the left compared to the one using the softplus feedback, implying that a shorter mitigation delay *τ* is sufficient to trigger oscillations. As for the softplus feedback, Arnold tongues emerge in the *τ* -*m*_max_-plane (Fig. 9b).

**Figure 8:**
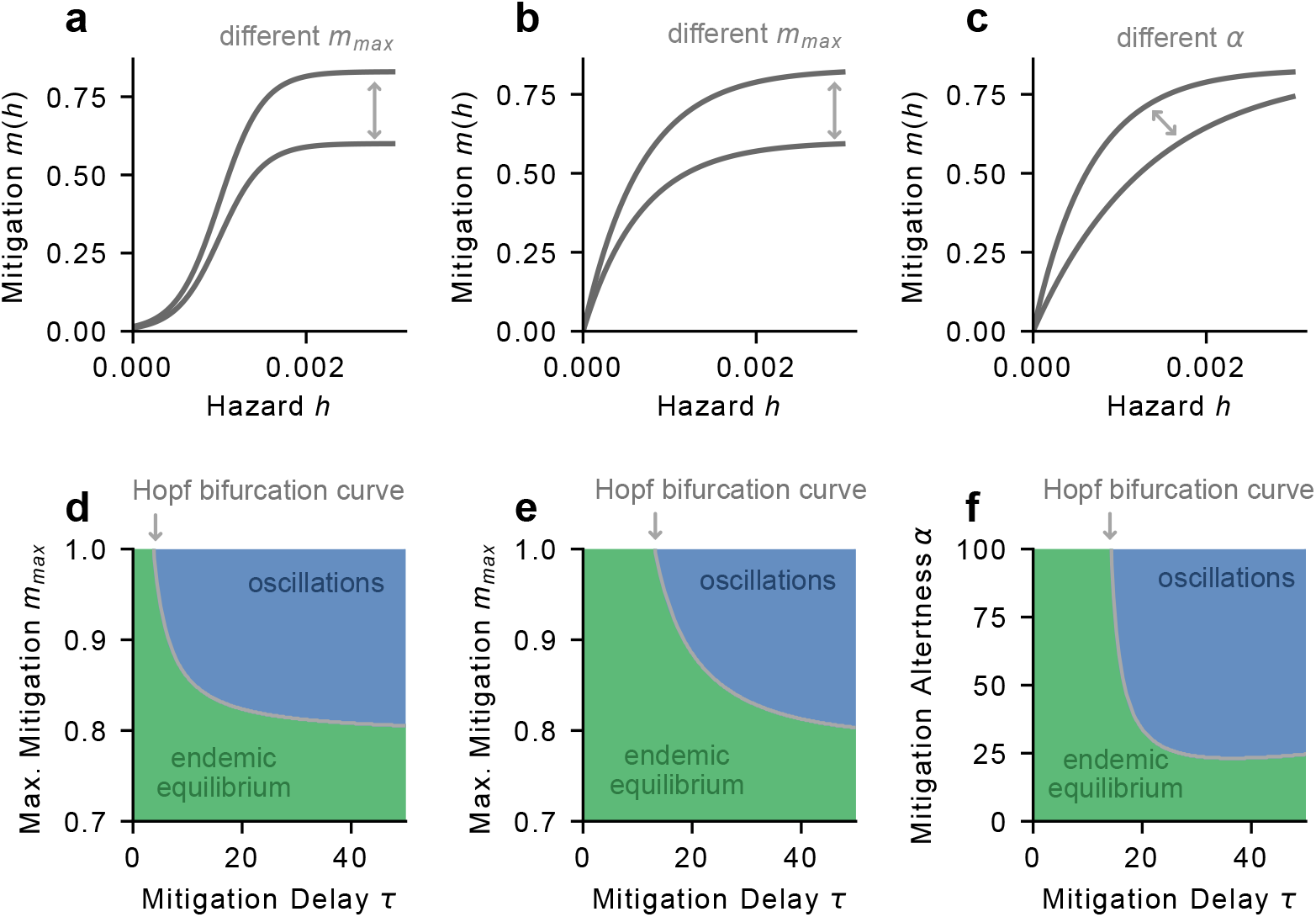
Various mitigation-hazard feedbacks trigger a Hopf bifurcation. The onset of oscillations due to a Hopf bifurcation is not unique to one specific example for the functional form of *m*(*h*). Instead, for different functional shapes of *m*(*h*) oscillations emerge when maximal mitigation *m*_max_ and mitigation delay *τ* are large enough (**a**,**d** for a logistic feedback, **b**,**e** for an exponential feedback). **c**,**f)** Mitigation alertness *α* is the other free variable of the exponential feedback. If it is large enough, namely mitigation increases fast enough with hazard, oscillations emerge. Model parameters used are found in Tab. 1. For **b**,**e** *α* = 1500 was used. For **c**,**f** *m*_max_ = 0.83 was used.

**Figure 9:**
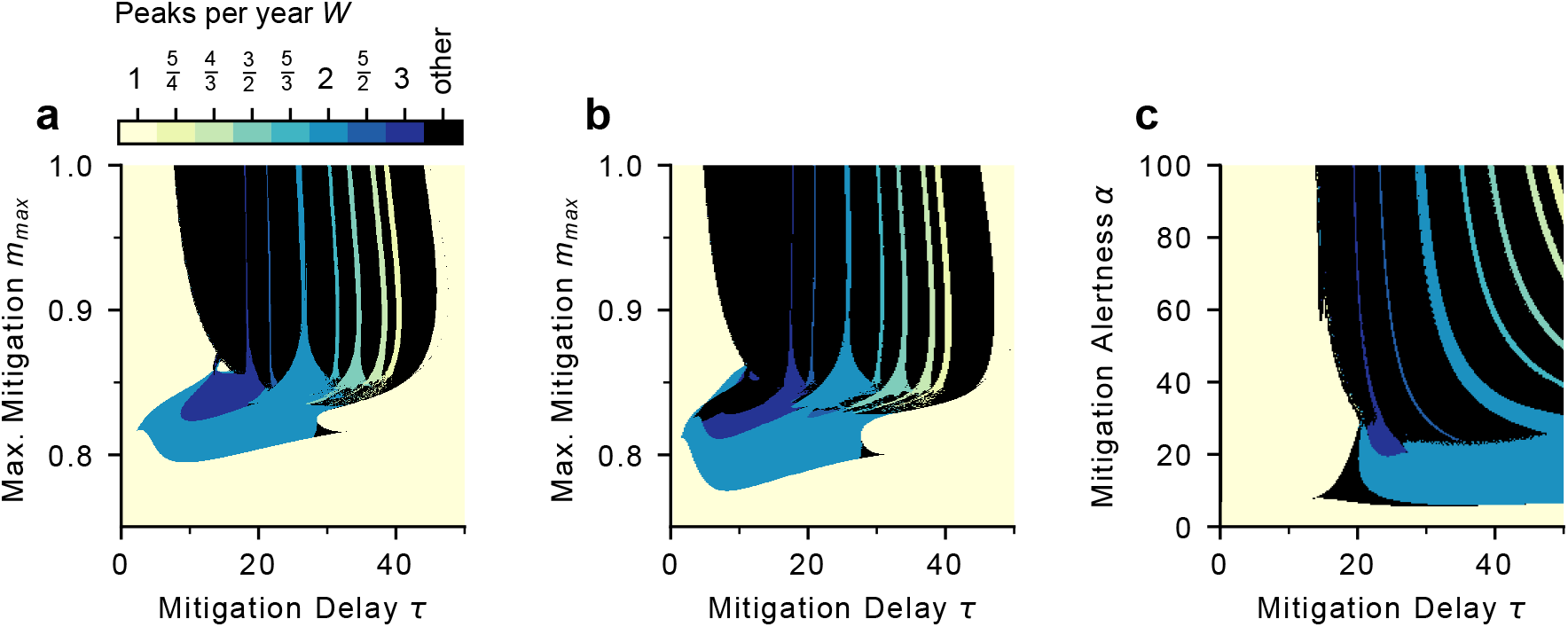
Arnold tongues emerge for different feedbacks *m*(*h*) (Fig. 8). **a)** Arnold tongues for the softplus feedback used in the main text. **b)** Arnold tongues for a logistic feedback (Eq. 21). **c)** The number of peaks per year *W* shows Arnold tongue-like structures using an exponential feedback (Eq. 22) and the mitigation alertness *α* as free parameter.

### Exponential feedback

The softplus as well as the logistic feedback are described by three parameters. In order to reduce the complexity of the model one could choose an exponential feedback with only two parameters:

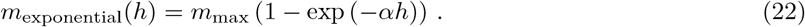

The exponent *α* describes the “mitigation alertness” of the society, i.e., how fast mitigation increases with hazard. It is thus comparable to the threshold hazard *h*_thres_ in the softplus feedback, as both scale the feedback in the *h*-direction. The stability diagram using mitigation alertness *α* as variable parameter shows that if it is large enough, i.e. the disease is mitigated fast enough, oscillations emerge (Fig. 8e,f). Furthermore, areas of synchronisation decrease in size as mitigation alertness *α* increases (Fig. 9c).

Varying the mitigation alertness *α* or the threshold hazard *h*_thres_ instead of the maximal mitigation *m*_max_ would thus pose a different way of looking at the onset of oscillations. However, throughout this manuscript we decided to consider *m*_max_ as a variable parameter since we believe that society decides how strongly it is willing to react at most rather than at what hazard level it does so. As the height of a wave is strongly influenced by the hazard level at which society reacts, this level could be imposed by e.g. hospital capacities, which is why we did not consider it as variable. The height of a wave is

All in all, the onset of oscillations around the endemic disease state seems to be a general feature if mitigation follows delayed infection numbers and is not unique to one specific type of feedback. As soon as the disease is mitigated strongly enough, namely with great strength at low infection numbers, oscillations emerge. Coupled with an external seasonal forcing, areas of phase-locking emerge that shrink in size with stronger mitigation, resulting in Arnold tongues.

## E Delayed feedback loop

The emergence of various complex dynamical regimes relies on the onset of oscillations through a Hopf bifurcation. This in turn does not only rely on the specification of a functional form for the mitigation *m*(*h*) but also on the kernel *K*(*t*) with which infection numbers are convoluted to yield the hazard *h*. We chose to use an Erlang kernel of second order 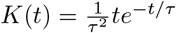 which has been shown to be in principle able to trigger oscillations [16]. By doing so, we assume that human risk perception only factors in past events. In other words, the hazard *h* only depends on infection numbers that have already happened but does not anticipate infections that are expected to come in the future.

In order to find evidence for such delayed risk perception during disease outbreak, we looked at Google Mobility Trends during the COVID-19 pandemic [41]. In particular, we looked at the category “retail and recreation”, which quantifies how mobility in public places such as restaurants, shopping centres, museums or movie theatres changed during the pandemic (compared to pre-COVID-19 times) as we believe that those provide a solid indicator for how threatened the society felt as a whole.

We found that for several waves of COVID-19 mobility changes followed the number of daily deaths with a delay. In other words, behavioural changes due to perceived risk followed the cause of the risk (here daily new deaths; infectious *I* in the model), providing evidence for delayed behavioural adaptation. For example, daily new deaths during the 2020 winter wave in Germany increased for several days to weeks before any significant change in mobility trends occurred (Fig. 10a). After a subsequent drop in mobility and a drop in daily new deaths, mobility increased again and reached the point where it initially started. This can be viewed as a hysteresis effect in clockwise direction: The input (daily new deaths) leads to two possible outputs (changes in mobility), depending on whether the input quantity increases or decreases. This effect can also be seen for waves in other countries such as the Netherlands (Fig. 10b). Analogously, in the SIRSm model a similar clock-wise hysteresis effect appears due to the delayed feedback mechanism, when plotting the normalised spreading rate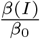 against the number of infectious *I* (Fig. 10c).

**Figure 10:**
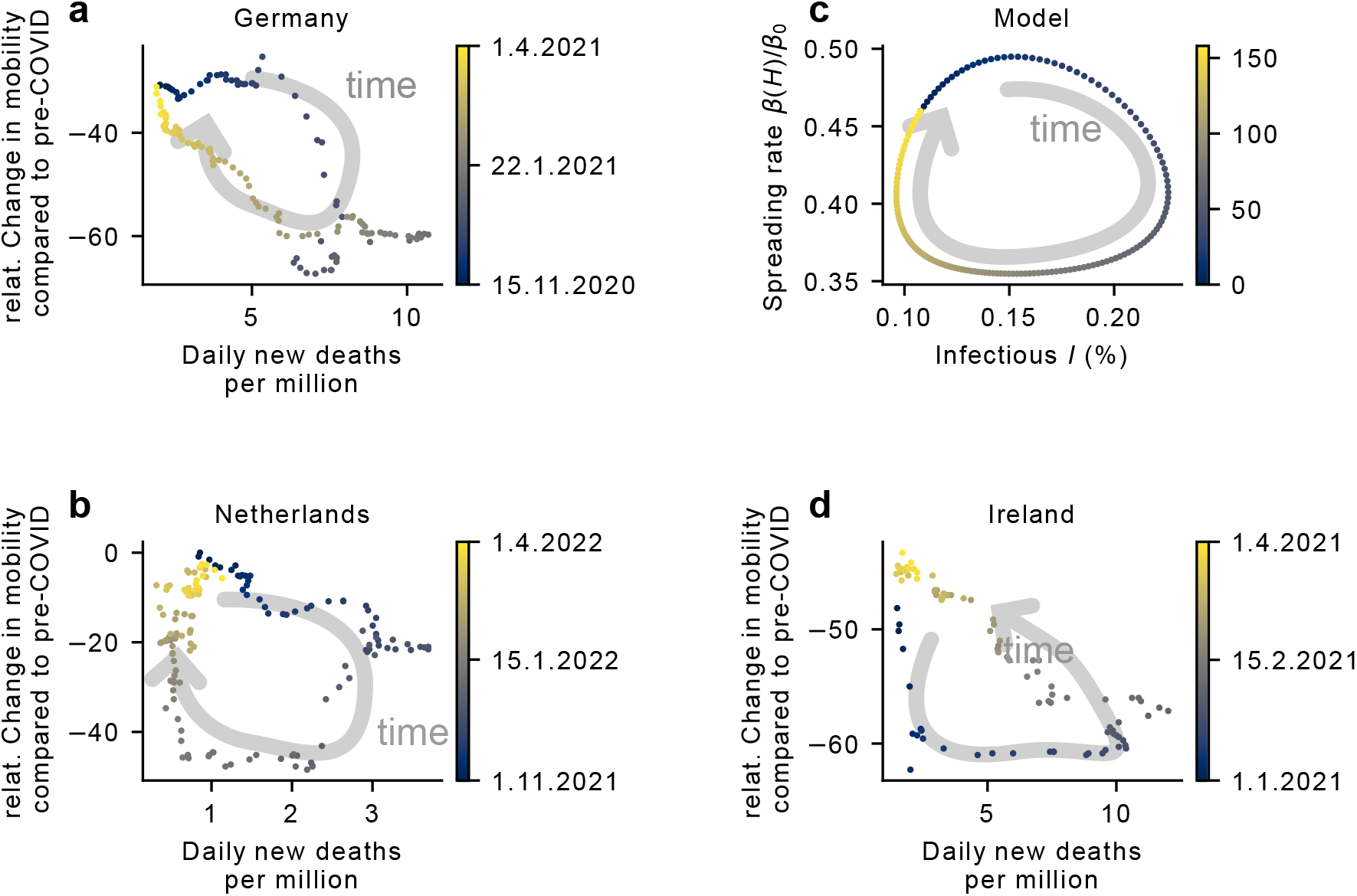
Google mobility trends provide evidence for delayed behavioural changes following infection numbers. **a**,**b)** Mobility in places such as restaurants and museums decreases (compared to pre-COVID-19) following increases in daily new deaths in Germany and the Netherlands during winter 2020 and 2021, respectively. However, mobility only adapts with a certain delay, resulting in clock-wise motion when plotting the change in mobility vs the daily new deaths: Deaths increase, which causes mobility to decrease due to perceived risk, which in turn reduces infections and thus the number of daily new death, causing an increase in mobility again. **c)** In the SIRSm model a delayed feedback mechanism between the incidence *I* and the spreading rate *β*(*H*) is implemented, also resulting in clock-wise motion in the *I*-*β*(*H*)-plane. **d)** Mobility changes in Ireland 2021 show an anticipation of rising infection numbers as mobility changes precede increasing daily new deaths, resulting in counter-clockwise motion. This is an effect not captured by the model.

While some waves do not show any clear relationship between mobility changes and daily new deaths, other waves also show an anticipation effect: Mobility changes in Ireland 2021 preceded an increase in daily new deaths, which lead to a hysteresis effect in counter-clockwise direction (Fig. 10d). One possible explanation is that neighbouring countries faced 2020/2021 winter waves at an earlier time and thus it was foreseeable that Ireland would soon face a similar outbreak, which lead to preemptive mitigation measures. The delayed feedback mechanism of the SIRSm model does not capture such anticipation of future infections, the implementation of which could be modelled by making the spreading rate dependent on the change of infections by writing 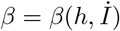 which we leave open for future research.

## F Different spreading rate

**Figure 11:**
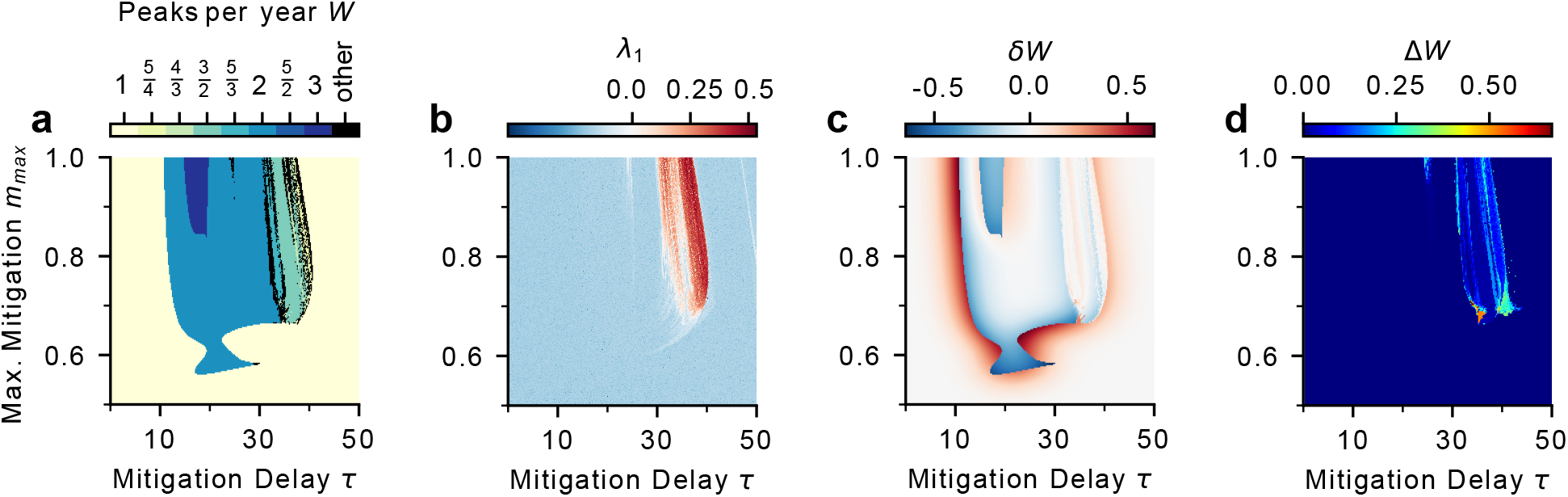
Complexity in the SIRSsm model also arises for a lower spreading rate. Using *R*_0_ = 2 different measures indicate complex dynamics in the *τ* -*m*_max_-plane. **a)** The number of peaks per year *W* does not show clear Arnold tongues as for *R*_0_ = 5, but neighbouring areas of different *W* interrupted by chaotic motion. **b)** The largest Lyapunov exponent *λ*_1_ can get similarly large as for *R*_0_ = 5, namely *λ*_1_ *≈* 0.5, implying significant divergence of initial conditions in about two years. **c)** Parameter variations obtained by Gaussian-weighted samples of trajectories around each point (with widths *σ*_*τ*_ = 4, 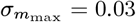) indicate the expected change in peaks per year *δW* as large as *δW* = 0.6. **d)** Coexisting attractors also emerge for *R*_0_ = 2 with different numbers of peaks per year as large as ∆*W* = 0.75 across attractors.

